# Oxytocin-augmented modular-based group intervention for loneliness: A proof-of-concept randomized-controlled trial

**DOI:** 10.1101/2023.10.30.23297746

**Authors:** Ruben Berger, Rene Hurlemann, Simone G. Shamay-Tsoory, Alisa Kantermann, Maura Brauser, Jessica Gorni, Maike Luhmann, Elisabeth Schramm, Johannes Schultz, Alexandra Philipsen, Jana Lieberz, Dirk Scheele

**Author notes:** **Corresponding authors:** Jana Lieberz: Department of Psychiatry and Psychotherapy University Hospital Bonn Venusberg-Campus 1 53127 Bonn, Germany Dirk Scheele Research Center One Health Ruhr of the University Alliance Ruhr Faculty of Medicine Ruhr-University Bochum 44780 Bochum, German. J.L. and D.S. contributed equally to this work (shared senior authorship).

## Abstract

**Introduction:** Loneliness poses a significant health problem and existing psychological interventions have shown only limited positive effects on loneliness. Based on preliminary evidence for impaired oxytocin signaling in trait-like loneliness, the current proof-of-concept study used a randomized, double-blind, placebo-controlled design to probe intranasal oxytocin (OT) as an adjunct to a short-term modular-based group intervention for individuals suffering from high trait-like loneliness (HL, UCLA loneliness scale ≥ 55).

**Methods:** Seventy-eight healthy HL adults (56 women) received five weekly group psychotherapy sessions targeting cognitive biases in loneliness. HL participants received OT or placebo before the intervention sessions. Primary outcomes were trait-like loneliness measured at baseline, after the intervention, and again at two follow-up time points (three weeks and three months), and, assessed at each session, state loneliness (visual analog scale), perceived stress (Perceived Stress Scale, PSS-10), quality of life (World Health Organization Five Well-Being Index, WHO-5), and the therapeutic relationship (Group Questionnaire, GQ-D).

**Results:** The psychological intervention was associated with significantly reduced perceived stress and improved trait-like loneliness across treatment groups, which was still evident at the 3-month follow-up. OT had no significant effect on trait-like loneliness, quality of life, or perceived stress. However, compared to placebo, OT significantly facilitated the decrease in state loneliness within sessions and significantly improved positive bonding between the group members.

**Conclusion:** Despite significantly improved trait-like loneliness after the intervention, OT did not significantly augment this effect. Further studies are needed to determine optimal intervention designs to translate the observed acute effects of OT into long-term benefits.

## Introduction

Loneliness is defined as an aversive emotional reaction, resulting from a perceived deficiency in the quality or quantity of social connections [1]. From an evolutionary standpoint, loneliness is postulated to be adaptive, since it serves as an aversive signal, which motivates people to re-connect, much like hunger motivates to eat [2, 3]. Persistent loneliness can trigger neurobiological and behavioral mechanisms with detrimental effects on mental and physical health [4–6]. Despite a close phenotypical overlap with depression and social anxiety, loneliness has been identified as a distinct construct [7, 8]. The prevalence of loneliness varies depending on assessment criteria and demographic factors such as age [9, 10], but loneliness has been increasingly recognized as an important sociopolitical and public health issue [11–13].

A recent meta-analysis showed that psychological interventions in general are effective in reducing loneliness [14]. Previous interventions included several different approaches, ranging from cognitive behavioral therapies to mindfulness-based interventions, and social skills training. Averaged across interventions, the meta-analysis yielded a small to medium effect size of g = 0.43, favoring the interventions against control conditions. This estimate, however, might be too optimistic as (1) there was no active control group in two thirds of the 31 analyzed studies, (2) only about 30% of the studies had a low risk of bias, and (3) the few larger studies showed smaller effect sizes than most studies with fewer participants. In addition, most studies had a very high attrition rate of up to 45% (mean 10.5%), most likely caused by long interventions (a mean of ten sessions of 1-2 h; duration ranging from two weeks to 12 months).

Considering the significant impact of loneliness for individuals and for society, there is an urgent need to develop more effective interventions [15]. A potential strategy is the pharmacological augmentation of psychological interventions via the hypothalamic peptide oxytocin (OT). OT is a key modulator of social cognition and behaviors including fear learning [16, 17], stress response [18, 19], parental and romantic bonding [20–23], empathy and social value representations [24, 25], and interpersonal trust [26–28]. Importantly, there is preliminary evidence that a chronic lack of social connections can interfere with OT-related social functions. In fact, reduced interpersonal trust has been linked to decreased oxytocinergic activity in response to positive social interactions in individuals with trait-like loneliness [29]. While previous studies yielded heterogenous findings about OT augmentation of psychotherapy [30, 31], a potential effect of OT as an adjunct to a modular-based intervention against loneliness has not been tested yet.

Thus, our randomized, double-blind, placebo-controlled trial investigated intranasal OT as a potential adjunct to a psychological short-term intervention for participants suffering from high trait-like loneliness (HL; UCLA loneliness scale ≥ 55; [32]). Based on the cognitive behavioral model of loneliness [4, 33], the short term group intervention consisting of five weekly sessions combined techniques from different evidence-based psychotherapies and targeted critical psychological mechanisms of trait-like loneliness, such as social hopelessness, distrust, limited social skills, and maladaptive thinking (dysfunctional cognitions).

Given the meta-analytic findings of psychological interventions on loneliness, the first of our preregistered hypotheses was that the group intervention would reduce trait-like loneliness (H1.1) and state loneliness (H1.2). We further expected that the psychological intervention would improve perceived stress (H2.1) and increase quality of life (H2.2). Our third hypothesis was that OT would facilitate the development of positive bonding as perceived by the participants (H3). Moreover, we hypothesized that OT would augment the positive effects of the intervention on trait-like loneliness (H4.1), state loneliness (H4.2), perceived stress (H4.3), and quality of life (H4.4). Finally, we tested the effects of the psychological intervention on secondary outcomes consisting of loneliness-related depressive symptoms and social anxiety. Moreover, we explored OT effects on the therapeutic relationship as seen by the therapist. Further exploratory analyses were conducted to examine possible long-term effects of the intervention with follow-up measurements three weeks and three months after the intervention. To further characterize the HL participants and to control for naturally occurring changes in the outcome measures, we included a control group involving healthy participants with low loneliness (LL) scores that did not receive the intervention.

## Material and methods

### Study design

The study design was registered at ClinicalTrials.gov (NCT04137432), and the analysis plan was pre-registered before conducting any analyses (https://osf.io/qpfw3). Healthy subjects suffering from trait-like loneliness (UCLA loneliness scale; UCLA-L scale [32, 34]; i.e., ≥ 55, i.e., at least 1.5 standard deviations above the mean score in the original validation study, cf. [32]) took part in the randomized, double-blind, parallel-group study design and received a modular-based group intervention to reduce loneliness. In half of the intervention groups, participants received 24 International Units (IU) 30 minutes before the group sessions and the other half received a placebo spray (PLC), that contained identical ingredients except for the peptide itself. There is strong evidence that intranasal OT reaches the brain [35] and 24 IU has been identified as the most effective dose to modulate amygdala reactivity in men [36]. Groups were randomized to each treatment condition with all participants of a group receiving the same treatment for all intervention sessions, that is, either OT or PLC. The allocation sequence was concealed from the therapists and the participants throughout the whole trial. As a control group, 49 healthy participants with low loneliness scores (UCLA-L scale ≤ 35) were included. The LL participants did not receive an intervention or treatment but were tested with the same psychometric measures as the HL participants at study entry and after a time interval comparable to the duration of the intervention of HL participants (see *Experimental procedure*). All participants (HL and LL) underwent functional magnetic resonance imaging (fMRI) before the start and after the end of the intervention (results will be presented elsewhere).

### Participants

We recruited participants via online advertisements and postings on social media platforms, a regional newspaper article, and physical flyers in healthcare as well as university facilities from June 2019 to November 2021. Interested individuals were asked to fill out an online questionnaire that included the UCLA-L scale and questions regarding the exclusion criteria (current psychiatric illness, current psychiatric medication or psychotherapy, MRI contraindication such as claustrophobia, and oxytocin contraindication such as cardiovascular diseases or pregnancy). A total of 167 HL individuals who fulfilled our inclusion criteria (UCLA-L score ≥ 55, aged 18-65 years) were invited to a first screening session (T0) which included an in-depth query of the exclusion criteria. The Mini International Neuropsychiatric Interview (MINI) [37] was conducted to rule out participants fulfilling the criteria for a psychiatric disorder. Included participants were then randomly allocated to the next possible intervention group. All participants who were assigned to an intervention group after finishing the first fMRI session (T1) and who participated at the first intervention session (T2) were included with respect to the intention-to-treat sample. In total, we enrolled 79 participants receiving either OT or PLC, resulting in 12 intervention groups. We excluded one participant from analyses because of concurrent participation in a psychotherapy. Thus, the final sample consisted of 43 HL participants in the OT group and 35 HL participants in the PLC group. The 49 LL participants had to fulfill the same inclusion and exclusion criteria as the HL participants, except for the UCLA-L score, which had to be under 35. For details of the enrollment process, see **Supplementary Figure S1**.

### Experimental procedure

All HL participants meeting our inclusion criteria evaluated through the online questionnaire were invited to a screening session (T0; see section *Participants*), during which psychiatric symptoms and social network characteristics were assessed. Following the screening session and before the start of the respective group intervention, participants completed further questionnaires during the first testing session (T1) to assess baseline trait-like loneliness and quality of life. The group intervention then started with a first introduction session (T2) and was carried out by one of two trained psychotherapists. The intervention was limited to five weekly sessions (including the first introduction session; T2).

Given the kinetics of central OT effects [36] the duration of each intervention session was limited to one hour. Notably, no OT was administered during T2 as previous findings indicated possible anxiogenic effects of OT in therapeutic situations involving strangers [31]. Therefore, the first group session allowed the participants to become familiar with the therapeutic context prior to the first OT administration. Due to recruiting difficulties during the Corona pandemic, we had to adapt our original study protocol such that the minimum group size was reduced to four individuals.

Our short-term group intervention included modules focusing on the critical psychological phenomena of long-lasting loneliness: such as social hopelessness, distrust, limited social skills, and dysfunctional cognition, leading to a lack of rewarding interpersonal relationships. As loneliness is not a psychiatric disorder, most psychotherapies have not been directed toward this problem specifically, and yet many psychotherapeutic techniques are well suited to address the behavioral, interpersonal, cognitive, and emotional correlates of loneliness. Utilizing techniques from different evidence-based psychotherapies matching the psychological correlates of loneliness, we developed a modular approach using tools mainly from Interpersonal Psychotherapy (IPT), Acceptance and Commitment Therapy (ACT), Cognitive Behavioral Analysis System of Psychotherapy (CBASP), and Social Skill Training (SKT). For details of the content of the psychological intervention see the Supplementary Information. After completing the intervention, participants filled out questionnaires regarding their loneliness, quality of life, and psychiatric symptomatology on a separate testing day (T7). Follow-up measurements of the aforementioned variables were further collected three weeks (T8) and three months (T9) after finishing the intervention. Participants of the LL group completed testing sessions T0, T1, and T7 in a comparable time interval without any treatment in between. A schematic overview of the study design for HL participants is shown in **Figure 1**.

**Fig. 1.**
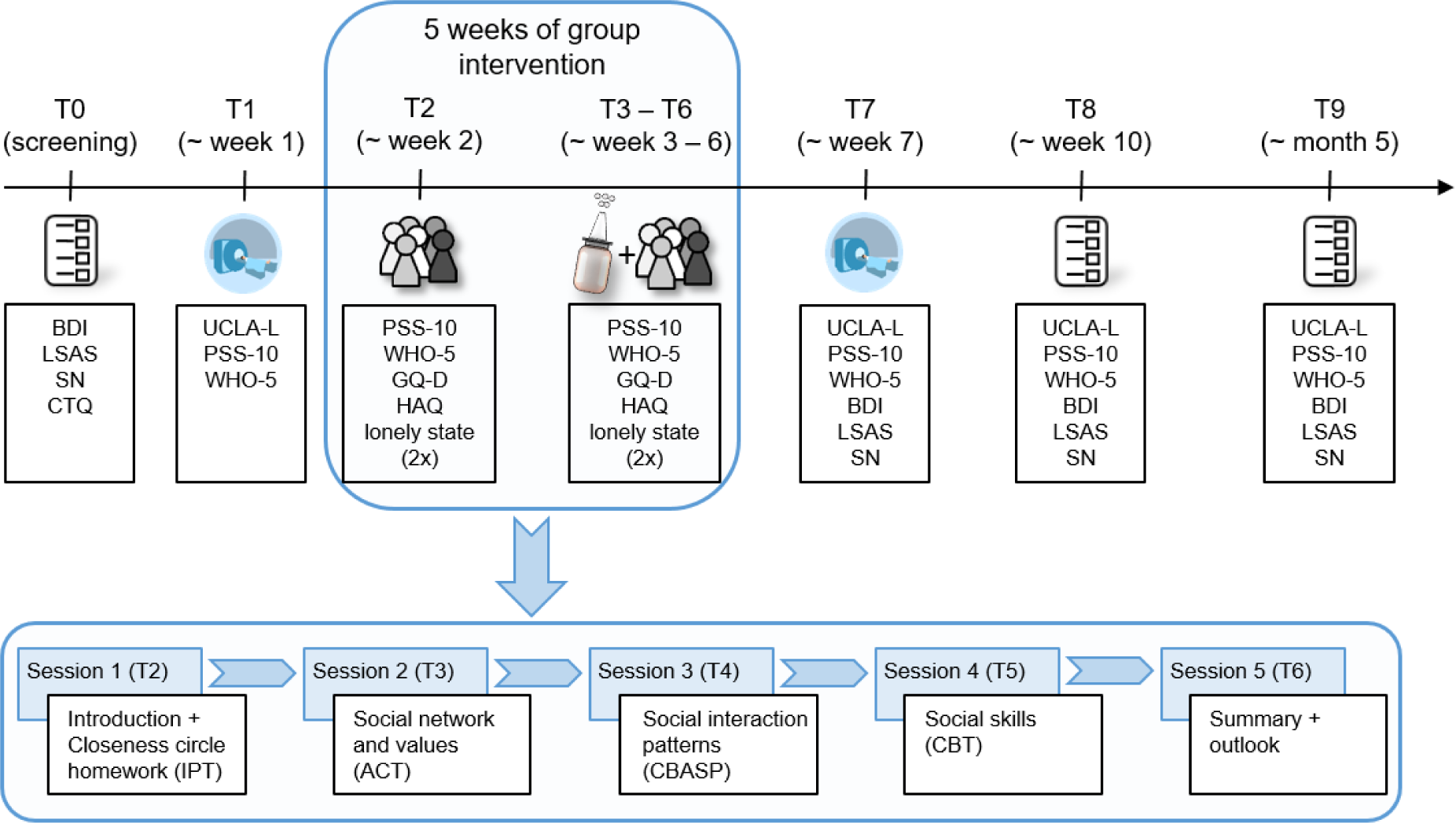
Illustration of the study design. The top row shows the time frame and collected measurements during the study. The second row depicts the five intervention sessions with an overview of the topics of each session. Abbreviations: ACT, Acceptance and Commitment Therapy; BDI, Beck’s Depression Inventory; CBASP, Cognitive Behavioral Analysis System of Psychotherapy; CBT, Cognitive Behavioral Therapy; CTQ, Childhood Trauma Questionnaire; GQ-D, Group Questionnaire; HAQ, Help Alliance Questionnaire; IPT, Interpersonal Psychotherapy; LSAS, Liebowitz Social Anxiety Scale; PSS-10, Perceived Stress Scale; SN, Social Network Index; T0-T9, testing session 0 (screening)-9; UCLA-L, UCLA Loneliness Scale; WHO-5, World Health Organization Five Well-Being Index.

### Measures

We measured trait-like loneliness using a validated German version of the revised UCLA-L scale with a 20-item 4-point Likert scale with scores ranging from 20 to 80 [32]. Specifically, the UCLA-L scale was used in an online questionnaire to identify participants who fulfilled our inclusion criteria (UCLA-L score of ≥ 55). Moreover, we assessed the UCLA-L scale at T1 (before the start of the intervention) and T7 (after the intervention) and at both follow-up time points three weeks (T8) and three months (T9) after the end of the intervention period. Furthermore, to assess state loneliness, participants rated their current feelings of loneliness on a Visual Analog Scale (VAS) from 0 (“not lonely at all”) to 100 (“very lonely”) from sessions T2 to T6 twice, i.e., at the beginning and at the end of each session.

Quality of life was assessed by 5 items of the World Health Organization Five Well-Being Index with a 5-point Likert scale with scores ranging from 0 to 25 (WHO-5; [38]) and subjective stress was measured by 10 items of the Perceived Stress Scale with a 5-point Likert scale with scores ranging from 0 to 40 (PSS-10; [39, 40]). Participants completed both questionnaires at the beginning of each testing session (T1 to T7) and at the follow-up measurements (T8 and T9). As participants were tested once a week during the group intervention period, instructions of the PSS-10 were changed for these measurements and referred to perceived stress in the last week instead of the last month (as assessed in the remaining testing sessions). To assess the therapeutic relationship towards the whole group, other individual participants, and the therapists, the three subscales of the Group Questionnaire GQ-D (30 items with a 7-point Likert scale) Positive Bonding (PB; range 4-28 for other participants and the therapist; range 5-35 for the whole group), Positive Working Relationship (PW; range 4-28 for the therapist and other participants), and Negative Relationships (NR; range 3-21 for the therapist, other participants, and the whole group) [41, 42] were completed by the group members at the end of each intervention session (T2 to T6). The perceived therapeutic alliance by the therapist was assessed at the end of each intervention session (T2 to T6) for exploratory purposes using the Help Alliance Questionnaire (12 item with a 6-point Likert scale) with scorers ranging from 12 to 72 (HAQ; [43]).

As secondary outcomes, depressive and social anxiety symptoms were assessed with the Beck Depression Inventory (21 items with a 4-point Likert scale) with scores ranging from 0 to 63 (BDI, [44]) and the Liebowitz Social Anxiety Scale (50 items with a 4-point Likert scale) with scores ranging from 0 to 72 (LSAS, [45]) at the screening session (T0), after finishing the intervention period (T7), and at both follow-up measurements (T8 and T9). For exploratory purposes and to characterize the sample in a more comprehensive way, we also assessed the social network size and diversity (i.e., number of social roles, number of people in one’s social network, and embedded networks) by using the social network index (SN; [46]). Like depressive symptomatology and social anxiety, the SN was completed at T0, T7, and at both follow-up measurements (T8 and T9). At the screening session (T0), the history of childhood maltreatment was assessed by the Childhood Trauma Questionnaire (CTQ; 25 items with a 5-point Liker scale with scores ranging from 25 to 125 [47]).

### Statistical analysis

We performed intention-to-treat analyses to avoid inflated type I-errors due to the inclusion of participants who did not complete the intervention. Intention-to-treat analyses provide a more real-world perspective on the efficacy, tolerability, and acceptability of an intervention. Therefore, all participants who received at least the first intervention session (T2) were considered for statistical analyses. To account for the multilevel, longitudinal data structure, we analyzed primary outcomes with linear mixed models with repeated measures.

To test our hypotheses, the following fixed-effects factors were included as predictors in their respective models:

1. To test the assumption that the psychological intervention has positive effects on feelings of loneliness (H1.1), testing session (T1, T7) was fitted as a fixed-effects factor to predict UCLA-L scores at both measurement occasions. For the model including state loneliness as the dependent variable (H1.2), both within-session time (start/end of intervention session) and testing session (T2, T3, T4, T5, T6) as well as their interaction were fitted as categorical fixed-effects factors. Similarly, both PSS-10 and WHO-5 scores were considered as dependent variables (H2.1 and H2.2). In these models, testing session (T1, T2, T3, T4, T5, T6, T7) was included as categorical fixed-effects factor.
2. With respect to OT effects on the therapeutic relationship as experienced by the participants (H3.1 and H3.2), the scores of the GQ-D subscales were the dependent variables and treatment (PLC, OT), testing session (T2, T3, T4, T5, T6), and the interaction of treatment with testing session were included in the models as categorical fixed-effects factors.
3. Likewise, treatment (PLC, OT) and its potential interactions with the respective fixed-effects factors as specified above were included as additional fixed-effects factors to investigate OT effects on loneliness, stress, and quality of life (H4.1 to H4.4).
4. For all analyses, sex and age (grand mean centered) assessed at T0 were included as covariates. To account for possible influences of the COVID-19 pandemic, an additional binary covariate was included in the models, which specified whether testing sessions were completed prior to the pandemic (i.e., in 2019 or the beginning of 2020) or during the pandemic.
5. To explore intervention and OT effects on the secondary outcomes, the same models as specified for the first hypothesis (H1.1 and H4.1, respectively) were used to predict BDI and LSAS scores, respectively.

For further details of the statistical analyses see the Supplementary Information.

## Results

### Study sample

HL individuals were characterized by increased psychopathological symptoms compared to the LL control group at study entry (for a comparison of sociodemographic data, see **Supplementary Table S1**). As such, lonely individuals reported increased depressive symptoms (*t*(108.4) = 10.24, *p* < 0.001, *d* = 1.57), social anxiety (*t*(117.7) = 8.78, *p* < 0.001, *d* = 1.38), and perceived social stress (*t*(100.9) = 10.88, *p* < 0.001, *d* = 1.98) in addition to decreased quality of life (*t*(119.8) = -8.73, *p* < 0.001, *d* = -1.51; see **Supplementary Table S2**). Moreover, as expected, HL participants reported more severe childhood maltreatment (*t*(124.8) = 6.55, *p* < 0.001, *d* = 1.07) and increased objective social isolation in form of smaller social networks (number of social roles: *t*(94.6), = -7.63, *p* < 0.001, *d* = -1.42; number of people: *t*(74.6) = -8.31, *p* < 0.001, *d* = -1.66; number of embedded networks: *t*(74) = -7.89, *p* < 0.001, *d* = -1.59). Importantly, however, within the HL group, participants receiving OT did not differ from those receiving PLC at study entry in either sociodemographic variables (see **Supplementary Table S3**), loneliness, or psychiatric symptomatology including depressive symptoms, social anxiety, childhood maltreatment, perceived stress, and quality of life (all *t* < 1.90, all *p* > 0.06; see **Supplementary Table S2**).

### Intervention effects

As expected, across all HL participants, the psychological intervention was associated with a reduction of trait-like loneliness as evident in significantly decreased UCLA-L scores from study entry at T1 to post intervention at T7 (H1.1) (*b* = -1.90, SE = 0.87, *t*(68) = -2.19, *p* = 0.03, *d* = -0.33; shown in **Fig. 2A**). Moreover, for state loneliness we found a significant interaction of testing sessions and within-session time (H1.2) (*F*(4,315.2) = 2.60, *p* = 0.04). Thus, changes in state feelings of loneliness from the start to the end of an intervention session significantly differed between testing sessions (T2 to T6). Specifically, post-hoc tests revealed an initial increase in state loneliness at T3, which differed significantly from the decrease observed in the following testing sessions (difference from pre to post at session T3 vs. T4: *b* = 11.24, SE = 4.14, *t*(316) = 2.72, *p* = 0.06, *d* = 0.31; T3 vs. T6: *b* = 11.60, SE = 4.12, *t*(313) = 2.82, *p* = 0.05, *d* = 0.29; all other comparisons of changes in pre-post values between testing sessions *t* < 2.00, *p* > 0.37).

**Fig. 2.**
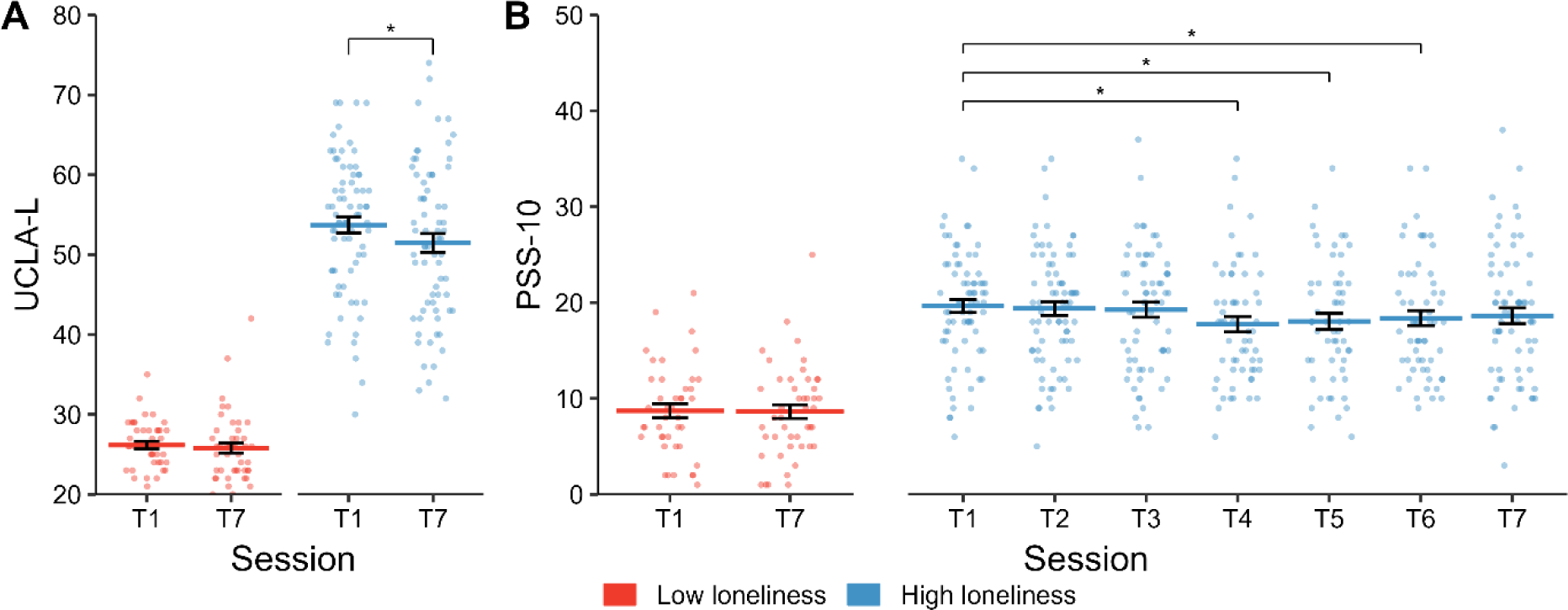
The group-based intervention was associated with a significant reduction of trait-like loneliness in individuals suffering from high loneliness (HL) from study entry at T1 to the post measurement at T7 (**A**). Likewise, the group-based intervention was linked to a significant decrease in perceived stress across sessions (T1-T7; **B**). Error bars indicate standard errors of the mean. Abbreviations: PSS-10, Perceived Stress Scale; T1-T7, testing session 1-7; UCLA-L, UCLA Loneliness Scale; \**p* < 0.05.

In addition to the observed reduction of loneliness and in line with hypothesis 2.1, perceived psychosocial stress decreased over time (main effect of testing session: *F*(6,373.5) = 2.62, *p* = 0.02, shown in **Fig. 2B** and **Supplementary Information**). However, no significant intervention effects were observed for quality of life (H2.2.; main effect session: *F*(6,387) = 0.69, *p* = 0.65), depressive symptomatology (*b* = 0.72, SE = 0.95, *t*(70.3) = 0.76, *p* = 0.45, *d* = 0.14), or social anxiety (*b* = 1.63, SE = 2.09, *t*(69.8) = 0.78, *p* = 0.44, *d* = 0.06). In contrast to the HL group, no significant changes in loneliness or perceived stress could be observed for the LL group between T1 and T7 (see **Supplementary Table S2**).

### Oxytocin effects

In line with our hypotheses, the OT treatment had a significant positive effect on the therapeutic relationship of HL participants (H3). Importantly, while a positive working relationship significantly improved from the start of the psychological intervention at T2 to the end at T6 across all participants (main effect of testing session: *F*(4,202) = 14.35, *p* < 0.001), OT specifically boosted positive bonding. In particular, positive bonding to the whole group increased significantly more in intervention groups receiving OT (interaction of treatment and testing session: *F*(4,234.5) = 2.83, *p* = 0.03; shown in **Fig. 3** and **Supplementary Information**). No further significant effects of treatment or testing session were observed for any of the GQ-D subscales (all further main effects of treatment or testing session and interactions of treatment and testing session: all *F* < 2.28, all *p* > 0.06).

**Fig. 3.**
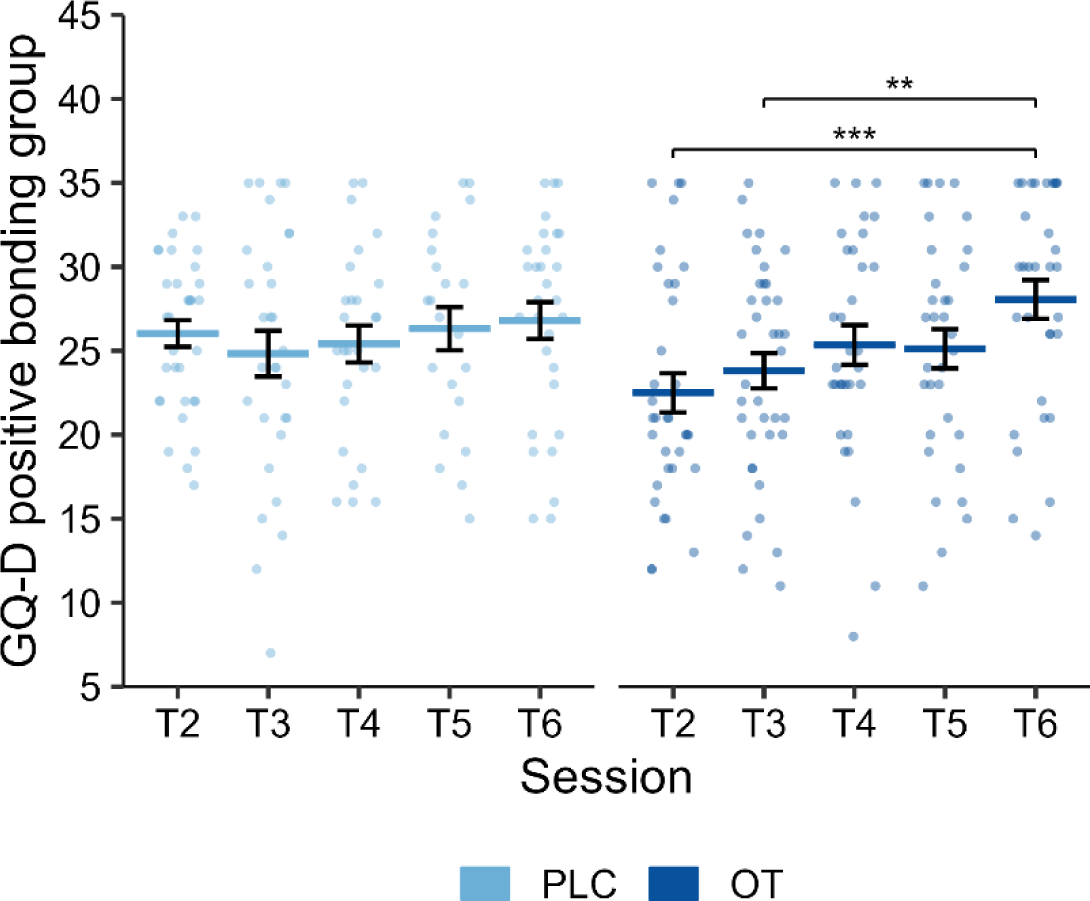
Positive bonding to the whole group increased significantly stronger in groups receiving intranasal oxytocin (T2-T6) compared to placebo. Error bars indicate standard errors of the mean. Abbreviations: GQ-D, Group Questionnaire; OT, oxytocin; PLC, placebo; T2-T6, testing session 2-6; \*\**p* < 0.01, \*\*\**p* < 0.001.

We did not observe a significant main or interaction effect of OT on trait-like loneliness (H4.1). Importantly, however, OT significantly improved the psychological intervention effects on state loneliness (H4.2) (interaction of treatment with within-session time: *F*(1,306.3) = 4.45, *p* = 0.04): Across intervention sessions, OT was associated with a decrease in state loneliness from the start of the testing session to the end (*b* = -4.62, SE = 1.78, *t*(305.6) = -2.60, *p* = 0.04, *d* = -0.34), whereas state loneliness did not change significantly within a testing session after PLC treatment (*b* = 1.02, SE = 2.00, *t*(306.8) = 0.51, *p* > 0.99, *d* = -0.10; shown in **Fig. 4**). Notably, further exploratory post-hoc tests indicated that OT prevented the observed increase in state loneliness at session T3 (see intervention effects), which was evident after PLC treatment (*b* = 14.16, SE = 4.30, *t*(309) = 3.29, *p* = 0.02, *d* = 0.48) but absent after OT treatment (*b* = -0.99, SE = 3.83, *t*(304) = -0.26, *p* > 0.99, *d* = -0.03). No further significant effects of OT were observed for our primary or secondary outcomes (main effects or interactions of treatment; all *F* < 4.42, all *p* > 0.06; see also **Supplementary Table S4**).

**Fig. 4.**
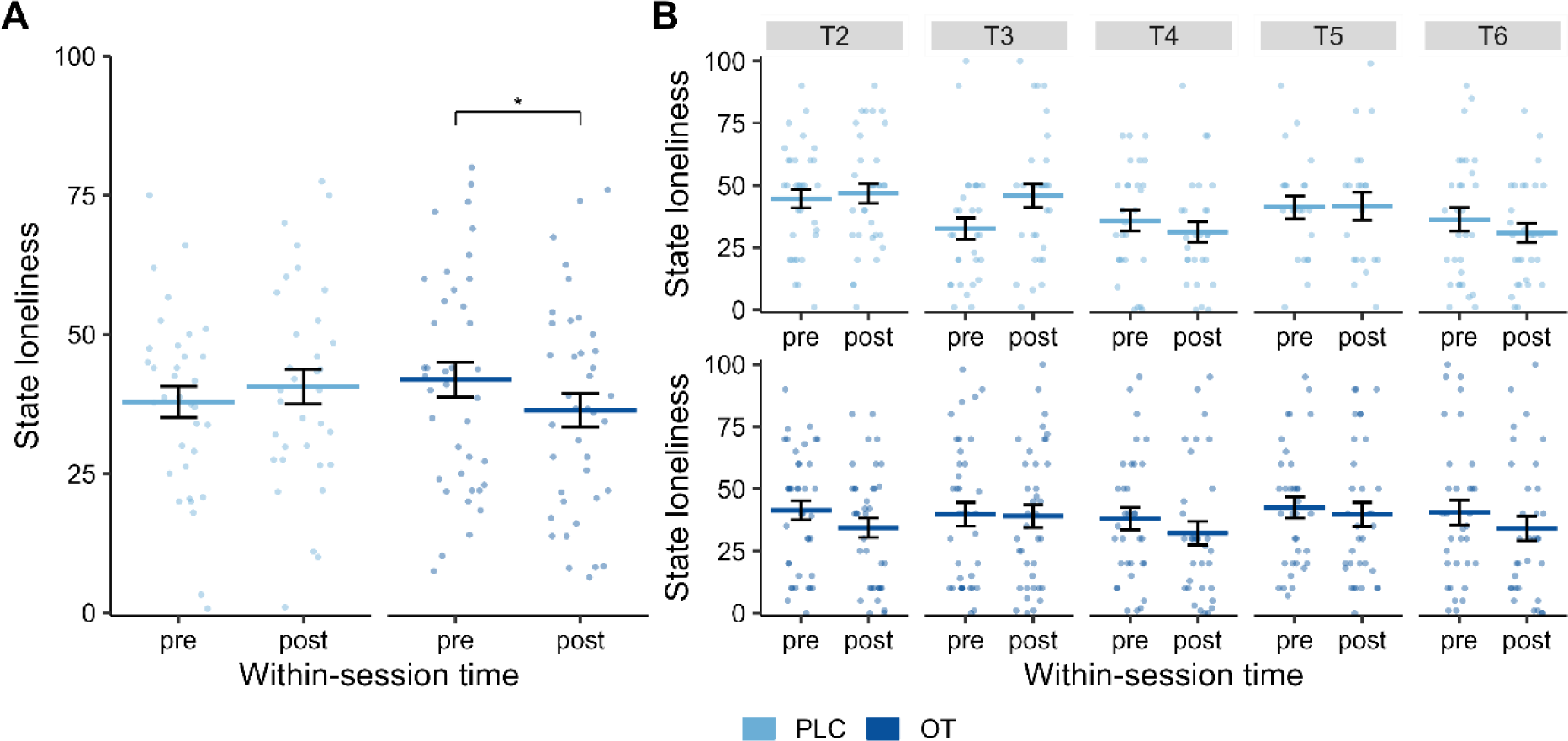
State loneliness was significantly lower after the intervention sessions compared to the start of the sessions in intervention groups with intranasal oxytocin treatment, but not placebo (**A**). In the placebo groups, state loneliness significantly increased at session T3 and this effect was absent in the oxytocin groups (**B**). Error bars indicate standard errors of the mean. Abbreviations: OT, oxytocin; PLC, placebo; T2-T6, testing session 2-6; \**p* < 0.05.

### Exploratory outcomes

In a first step, we explored whether positive effects of the OT treatment on the therapeutic relationship were also evident in an improved relationship as experienced by the therapists. In fact, the therapeutic relationship measured by the HAQ showed a descriptively overall better result in OT groups compared to the PLC groups (mean ± SD for OT: 46.70 ± 3.48; PLC: 44.27 ± 3.23). This pattern of results was observed for both HAQ subscales (relationship to the patients subscale for OT: 27.10 ± 2.08; PLC: 26.12 ± 1.93; satisfaction with the therapeutic outcome subscale for OT: 19.60 ± 1.61; PLC: 18.15 ± 1.56).

We then explored whether positive intervention effects on UCLA-L scores and perceived psychosocial stress were still evident in our follow-up measurements at three weeks (T8) and three months (T9) after completing the intervention. Importantly, across all HL participants, trait-like loneliness scores at T8 and T9 did not differ from those collected after finishing the intervention at T7 (*p* = 0.77) indicating a long-lasting decrease in loneliness. Along these lines, loneliness at T9 was still significantly decreased compared to study entry at T1 (*b* = -3.10, SE = 0.98, *t*(186) = 3.15, *p* = 0.01, *d* = -0.41; all other post-hoc comparisons: *t* < 2.14, *p* > 0.13). In contrast, no significant long-term effects were observed for perceived psychosocial stress (all post-hoc comparisons: *t* < 2.13, *p* > 0.16). However, a significant main effect of testing session after including T8 and T9 in the models indicated a delayed intervention effect on social anxiety (*F*(3,188.7) = 4.22, *p* = 0.006). Post-hoc tests revealed a significant reduction of social anxiety from study entry at T0 as well as from T7 after finishing the intervention to the first follow-up measurement at T8 (T8 vs. T0: *b* = -6.28, SE = 2.46, *t*(189) = - 2.55, *p* = 0.046, *d* = -0.38; T8 vs. T7: *b* = -7.82, SE = 2.48, *t*(187) = -3.15, *p* = 0.01, *d* = -0.45; all other post-hoc comparisons: *t* < 2.20, *p* > 0.08). No further significant delayed effects of the intervention were observed for stress, quality of life, or depressive symptomatology (all main effects of testing session: *F* < 0.90, *p* > 0.52). Likewise, OT treatment had no significant effects on our primary or secondary outcomes at the follow-up measurements at T8 and T9 (all interactions of treatment with testing session after including T8 and T9 in the models: *F* < 2.14, *p* > 0.09; see also **Supplementary Table S5**).

Finally, we explored whether the psychological intervention had positive effects on objective social isolation (i.e., social network sizes) in addition to the reported effects on loneliness. Indeed, the number of social roles of the participants and the number of people within one’s network increased from study entry to T7 (number of roles: *b* = 0.45, SE = 0.17, *t*(70.3) = 2.61, *p* = 0.01, *d* = 0.37; number of people: *b* = 1.20, SE = 0.61, *t*(70.2) = 1.96, *p* = 0.05, *d* = 0.31), whereas the number of embedded networks within one’s social network did not significantly change (*b* = 0.18, SE = 0.12, *t*(70.6) = 1.50, *p* = 0.14, *d* = 0.24). Changes within the social networks were independent of OT (all main effects or interactions with treatment: *F* < 1.42, *p* > 0.23; see **Supplementary Tables S2 and S5**).

For all models investigating OT effects, an interaction of treatment with sex was considered to test for potential moderations of treatment effects by sex. However, no interactions between sex and treatment were found in any of the models except for a sex-dependent change in social anxiety from study entry at T0 to T7 (interaction of sex, treatment, and session: *F*(1,65.4) = 4.83, *P* = 0.03; all other *F* < 2.11, all *p* > 0.15). Separate models for each sex indicated positive effects of OT on social anxiety in male participants (interaction of treatment with session: *F*(1,16.1) = 7.79, *p* = 0.01) that were absent in female participants (*F*(1,49.5) = 0.07, *p* = 0.78, for further information see supplementary results).

## Discussion

This study aimed to examine the effects of OT as an adjunct to a short-term psychological group intervention against loneliness. The psychological group intervention was associated with a small, but significant improvement of trait-like loneliness and psychosocial stress, and the former effect was still evident three months after the end of the intervention. Contrary to our hypothesis, OT had no further augmenting effect on these outcomes. Consistent with our hypothesis, however, OT significantly decreased state loneliness scores from the start to the finish of each group session, an effect not seen in PLC groups. Also consistent with our hypothesis, OT significantly improved positive bonding to the whole group. Furthermore, neither the intervention nor the OT treatment significantly changed quality of life or depressive symptoms. Interestingly, however, exploratory analyses revealed a delayed intervention effect of reduced social anxiety three weeks and three months after the group sessions which was further enhanced by OT in men.

Intriguingly, OT significantly facilitated the decrease in state loneliness within the sessions, but the peptide produced no significant effect on trait-like loneliness. More specifically, the deterioration of state loneliness during T3 in the PLC groups was prevented by OT. This second group session included psycheducation regarding the negative consequences and the vicious circle of development and maintenance of trait-like loneliness. In the following sessions there was a consistent within-session improvement of state loneliness. These following sessions focused more on symptom management and skill learning, possibly more likely to engender hope and relief. It is well known from different process-monitored psychotherapies that an initial problem activation may cause a temporary exacerbation of symptoms [48, 49]. These results provide further support for the notion that social effects of OT vary substantially between contexts [50, 51, 25]. Because of the observation that the adjunct application of 40 IU of OT before the first psychotherapy session resulted in acute anxiogenesis in patients with depression [31], we implemented a first meet-and-greet session without OT to create a safe environment. Nevertheless, OT did not significantly influence trait-like loneliness. Because significant effects of OT on positive bonding were evident in later sessions, it is conceivable that more sessions are needed for positive group dynamics to impact the therapeutic group process itself and, thereby, trait-like loneliness. Likewise, we did not observe a significant intervention effect on subjective well-being although there is a close link between loneliness and quality of life [52]. Along these lines, a longer intervention or a longer follow-up period may be necessary for changes to become evident. The intervention had no significant effect on social anxiety in the days following the intervention (at T7), but both the fear and avoidance of social situations were significantly reduced three weeks (T8) and three months (T9) after the intervention. Participants may need more time to learn and to practice new skills and behaviors in daily life and thereby reduce avoidance and anxiety through new experiences (confrontation) and cognitive change, both in the PLC and OT group. In line with this idea, a delayed reduction of anxiety has also been observed for group psychotherapy for major depressive disorder [53]. Interestingly, OT significantly enhanced intervention effects on social anxiety only in men, while the treatment effect on positive bonding was evident in both sexes. Sex-specific effects of OT have been observed in various domains [54–57] and appear to be especially pronounced for amygdala-related fear processing [58, 59]. By contrast, OT may exert similar effects on bonding-related reward processing in women and men [21, 60]. However, the current study was not designed to detect sex-specific OT effects and therefore these findings need to be interpreted with caution.

The OT-induced increase in positive bonding with the whole group is consistent with previous studies showing a significant OT effect on interpersonal trust [61, 26, 62], at least in participants with a low disposition to trust [27] like trait-like lonely individuals [29]. Mechanistically, OT may have influenced the biobehavioral synchrony between participants given previous findings that trait-like loneliness is associated with reduced social synchrony [63], and that OT can improve synchrony in group psychotherapies [64]. OT-based synchrony predicted treatment outcomes in a dyadic psychotherapy for depression [65], but an increase in OT levels during psychodynamic treatment sessions correlated with more instances of conflict and rupture in the alliance with the therapist [66]. We did not measure OT levels during the sessions in the present study, but improved bonding after exogenously elevated OT levels suggest that OT effects differ between cognitive-behavioral and psychodynamic interventions or might reflect differences between patients with depression and trait-like lonely individuals. Furthermore, the fact that the therapists also rated the therapeutic relationship more positive in OT groups than PLC groups indicates that the OT-related bonding effects could be perceived by the therapists.

The findings of the present study need to be considered in the context of the following limitations: The lack of a sham intervention or waiting group hampers any conclusions about the efficacy of the group-based psychological intervention. The significant, but relatively small decrease in trait-like loneliness after the psychological intervention is not surprising considering the short-term design in the current study. The intervention consisted of five sessions that each were limited to one hour to account for the pharmacodynamic effects of OT [36]. Furthermore, the psychological intervention only transiently increased social network size and diversity, while the reduction of trait-like loneliness persisted even three months later. As such, the psychological intervention effect on loneliness cannot be explained as a byproduct of altered objective social isolation. Future studies with ecological momentary assessments are warranted to probe how intervention-related behavioral changes are implemented in real life.

Collectively, our results show that OT as an adjunct to a short-term group-based psychological intervention is feasible and has a positive effect in reducing state loneliness and in enhancing positive group relationships. However, we did not observe a significant reduction of trait loneliness. As loneliness is one of the strongest social determinants of somatic and mental health [13], further research is warranted to determine how significant augmentation effects of OT on acute loneliness can be intensified, translated, and carried forward to long-term benefits to improve trait-like loneliness.

## Statement of Ethics

The study was approved by the institutional review board of the Medical Faculty of the University of Bonn and conducted in accordance with the latest revision of the Declaration of Helsinki. Participants provided written informed consent after receiving a complete description of the study.

## Author contributions

R.B., J.L. and D.S. designed the experiment; R.B., M.B., J.G., and J.L. conducted the experiments; R.B., J.L., and D.S. analyzed the data. All authors wrote the manuscript. All authors read and approved the manuscript in its current version.

## Data availability statement

The data that support the findings of the present study are openly available in the repository of the Open Science Foundation at https://osf.io/kdy4p/.

