## Supplementary Information for "Oxytocin-augmented modular-based group intervention for loneliness: A proof-of-concept randomized-controlled trial"

### Supplementary material and methods

*The design of the five intervention sessions*

During the first intervention session (T2), the focus was set on an initial contact between the participants and a familiarization with the therapeutic group setting. After an initial introduction of the therapist, the participants were encouraged to introduce themselves and share with the group what motivated them to take part in the intervention and how loneliness plays a role in their lives. This should increase group coherence and reduce social anxiety towards the unfamiliar situation. Moreover, an overview of the agenda of the group sessions was given by the therapist to further reduce uncertainty and anxiousness of the participants regarding the intervention. At the end of the session, the participants were given homework consisting of exercises and worksheets which should be completed between every session. The procedure of providing homework after each session was chosen given the limited time during sessions due to the kinetics of central intranasal oxytocin (OT) effects [1]. Nevertheless, the therapeutic homework also left more room during the sessions for an exchange between group members, aiming to potentially further promote the positive augmentation effects of oxytocin. As a first homework, the participants were asked to fill in the so called “closeness circle”, used during Interpersonal Therapy (IPT) to visually assess the quantity and quality of their currant social network [2]. This visualization of the status quo of the social situation aimed at helping the participants to develop a better understanding of their individual cause of loneliness, applying the psychotherapeutic principals of clarification of meaning, problem actuation, and change motivation.

At the beginning of the second intervention session (T3), participants got the chance to ask questions and share their experiences regarding the “closeness circle”. With the aim of psychoeducation, the dangers and negative consequences of loneliness as well as a model developed by us describing the vicious circle of development and maintenance of loneliness were then introduced and discussed. The model describes how dysfunctional cognitions and behaviors can cause feelings of loneliness and how they intensify them. The psychoeducation followed the therapeutic strategy of guided discovery fostering an active participation of the group members. Moreover, participants were introduced to the concept of value-based living as part of the Acceptance and Commitment Therapy (ACT) [3]. This should help showcase where loneliness and associated avoidance inhibit value-consistent behavior and motivate participants to engage in cognitive and behavioral change towards their interpersonal goals. Therefore, participants were asked to perform an ACT exercise called “value-compass” as the second homework, which was developed to help identify and visualize personal values, associated personal goals, and (loneliness-related) barriers hindering a progression towards those personal value-based goals [4].

The third intervention session (T4) marked the transition from an initial phase of understanding to a more active phase focusing on cognitive and behavioral change. In the beginning of this session, the participants got the chance to exchange and discuss the homework, i.e., their personal values, resulting goals, and loneliness-related barriers by means of the “value-compass”. Subsequently, during the second half of the session, the Kiesler-model derived from the Cognitive Behavioral Analysis System of Psychotherapy (CBASP) was introduced to the participants. This model aims at explaining and predicting the causal relationship between one’s own behavior and the reactions of others, enabling behavioral change towards personal goals [5]. By providing insights and behavioral alternatives, this model should help the participants to break the self-fulfilling vicious circle of loneliness. To encourage exchange and collectively find more functional behaviors in different social situations, participants discussed problems related to social interaction and gathered adequate behavioral solutions. This was further facilitated by the therapist providing information from social skills training and the Kiesler-model, following the therapeutic change mechanism of mastery and coping as well as clarification of meaning [6].

In the beginning of the fourth intervention session (T5), participants were asked to discuss experiences from the last week, where they could implement (cognitive or behavioral) change, based on what they learned during the third session. Afterwards, the session focused on loneliness-related problems surrounding romantic relationships. Discussing both the feeling of loneliness caused by the lack of a relationship or elicited during a relationship was used as relatable vehicle to illustrate the differences and similarities of loneliness and isolation. Necessary social skills for finding a romantic partner as well as finding satisfaction within a relationship through speaking up about personal needs and demarcation were discussed within the group. This was supplemented by the therapist providing the group with fitting elements from social skills training. Finally, as homework, participants were encouraged to revisit their goals from T3 and try to apply new strategies they learned during the last sessions to approach those goals, applying the principal of resource activation [7].

In the fifth and final intervention session (T6), participants were encouraged to exchange experiences and take-home-messages most important to them over the course of the intervention. Also, the goals defined during T3 in the context of values were discussed. Participants were encouraged by the therapist to share which aspects of the intervention helped them the most to approach their value-based goals and what they want to further implement in their life. This should help generalize and consolidate new knowledge, skills, and social trust acquired during the intervention. In the end of the session, all participants got the chance to provide feedback about their experience during the group intervention.

*Statistical analysis*

Mixed models with repeated measures (MMRM) were calculated using the lme4 [8], lmerTest [9], and emmeans [10] package of the statistical software program R. All models were fitted using restricted maximum-likelihood and participants (level 2 unit) nested in intervention groups (level 3 unit) were included as random effects factors, allowing intercepts to vary across participants and intervention groups. When participants with low loneliness scores (LL) were included in the analysis (i.e., comparisons of baseline and post-intervention characteristics of the HL group with the LL group), each LL participant was treated as single level 3 unit (cf. [11]). To identify the maximal random effects structure supported by the data (cf. [12-14]), we implemented a stepwise approach. We started with an empty model without fixed effects factors to estimate the intraclass correlation coefficients. We then added the above specified fixed effects factors with fixed slopes to the model and successively increased the random effects structure for each model by allowing slopes to vary across participants and intervention groups. If the model fit did not improve significantly by adding a further random slope as indicated by deviance tests or if a model failed to converge after including the additional random slope, the respective random slope was excluded for the final model. The most complex model supported by the data was used to evaluate the significance of the fixed effects. After identifying the maximal random effects structure, covariates were included in the model and omnibus tests of main effects or interaction effects of fixed effects factors were tested by the ANOVA function of the lmerTest package. Post-hoc tests were calculated using pairwise comparisons as implemented in the emmeans package. To compute denominator degrees of freedom and *F*-statistics, Kenward-Roger's method was used. For all effects, *p*-values < 0.05 (two-tailed) were considered significant. *p*-values of post-hoc comparisons were adjusted for multiple comparisons using the Bonferroni-Holm method.

Our sample size is based on an a-priori power analysis. To the best of our knowledge, potential OT effects as an adjunct to group interventions for loneliness have not yet been investigated. However, previous studies have already investigated the effects of different kinds of interventions on the experience of loneliness. G*Power 3 [15] was used to determine the sample size which was needed to replicate previously reported intervention effects on loneliness. Specifically, for intervention effects on loneliness assessed by the UCLA-L scale in a pre-post design, a meta-analysis revealed a mean effect size of d = -0.49 [16]. To reliably detect an intervention effect of this size with a power of 0.95 (α = 0.05, two-sided, paired t-test), a total sample of 55 participants was needed. Assuming a drop-out rate of ~10%, we planned to include at least 60 individuals. Notably, our hypothesis of positive OT effects on the therapeutic relationship was based on previous findings showing that OT has positive effects on interpersonal trust [17] at least in participants with a low disposition to trust [18]. We thus additionally examined whether the planned sample size of 60 participants would be sufficient to detect OT effects on interpersonal trust as previously observed. Baumgartner et al. [17] reported an effect of OT of at least d = 0.78 on the neural processing of trust which was accompanied by behavioral OT effects. The power analysis (α = 0.05, two-sided, two-sample t-test) revealed that we would achieve a power of 0.85 to detect an effect of this size by including 30 participants per group, indicating that our planned sample size would also have an acceptable power to detect OT effects as reported previously.

**Supplementary results**

*Intervention effects*

We observed no significant differences in state feelings of loneliness depending on the testing sessions in general (main effect of session: $F(4,63.9) = 2.08$, $p = 0.09$) or within-session time (main effect of within-session time: $F(1,315.3) = 2.21$, $p = 0.14$) in general.

Compared to perceived stress scores at study entry, perceived psychosocial stress was significantly reduced from T4 on until the last intervention session at T6 (T4 vs. T1: $b = -1.98$, SE = 0.63, $t(374) = -3.16$, $p = 0.01$, $d = -0.31$; T5 vs. T1: $b = -1.76$, SE = 0.63, $t(373) = -2.79$, $p = 0.03$, $d = -0.25$; T6 vs. T1: $b = -1.60$, SE = 0.63, $t(374) = -2.56$, $p = 0.04$, $d = -0.18$; all other comparisons with T1: all $t < 2.06$, all $p > 0.12$).

*Oxytocin effects*

Successful blinding of treatment was confirmed by chi-squared tests with the participants' guesses of the received treatment being independent of the actual treatment ($\chi^2(2) = 0.82$, $p = 0.66$). The percentage correct estimates of treatment was even lower than chance (correct estimates: 33.33 %, $\chi^2(1) = 7.00$, $p = .008$).

Post-hoc tests with regard to positive bonding (PB) revealed significantly enhanced PB scores at T6 compared to T2 ($b = 5.58$, SE = 1.11, $t(234.3) = 5.02$, $p < 0.001$, $d = 0.95$) and T3 ($b = 4.54$, SE = 1.09, $t(235) = 4.17$, $p = 0.001$, $d = 0.58$) after OT treatment which was not evident for the placebo (PLC) treatment (T6 vs. T2: $b = 0.52$, SE = 1.13, $t(233) = 0.45$, $p > 0.99$, $d = 0.14$; T6 vs. T3: $b = 1.17$, SE = 1.17, $t(234.5) = 1.00$, $p > 0.99$, $d = 0.19$). Interestingly, the positive effect of OT seemed to be specific for positive bonding to the whole group as the scores of other PB subscales increased significantly over the time course of the psychological intervention (main effect of testing session for PB total score and further PB subscales: all $F > 6.57$, all $p < 0.001$) but were not affected by treatment (all treatment x testing session interactions: $F < 2.13$, $p > 0.07$).

*Exploratory outcomes – long-term effects of the psychological intervention*

Trait-like loneliness scores at T8 and T9 did not differ from those collected after finishing the intervention at T7 (T8 vs. T7: $b$ = 0.99, SE = 0.89, $t$(184) = 1.12, $p$ = 0.77, $d$ = 0.26; T9 vs. T7: $b$ = -1.11, SE = 0.98, $t$(185) = -1.14, $p$ = 0.77, $d$ = -0.13). Notably, a delayed intervention effect was evident for both Liebowitz Social Anxiety Scale (LSAS) subscales (main effect of testing session for the anxiety subscale: $F$(3,189.1) = 2.78, $p$ = 0.04; avoidance subscale: $F$(3,188.8) = 5.13, $p$ = 0.002) but more pronounced for the avoidance subscale (T8 vs. T0: $b$ = -4.93, SE = 1.42, $t$(189) = -3.47, $p$ = 0.003, $d$ = -0.44; T8 vs. T7: $b$ = -4.38, SE = 1.44, $t$(187) = -3.05, $p$ = 0.01, $d$ = -0.44; all other post-hoc comparisons: all $t$ < 2.09, all $p$ > 0.11) than for the anxiety subscale (T8 vs. T7: $b$ = -3.45, SE = 1.32, $t$(187) = -2.62, $p$ = 0.048, $d$ = -0.34; all other post-hoc comparisons: all $t$ < 2.28, all $p$ > 0.09).

Briefly after finishing the intervention (T7), the number of roles and people within one's network decreased and changes were no longer observed at both follow-up measurements T8 and T9 (all post-hoc comparisons: all $t$ < 2.41, all $p$ > 0.08; main effect of testing session for number of embedded networks after including T8 and T9 in the model: $F$(3,189.5) = 1.58, $p$ = 0.20).

*Sex-dependent oxytocin effects on social anxiety*

Separate models for each sex revealed a significant OT effect in male participants (interaction of treatment with session: $F$(1,16.1) = 7.79, $p$ = 0.01) that was absent in female participants ($F$(1,49.5) = 0.08, $p$ = 0.78). Further post-hoc tests in the male sample indicated a decrease in scores of the LSAS from T0 to T7 after OT ($b$ = -8.07, SE = 4.57, $t$(16.3) = -1.76, $p$ = 0.29, $d$ = -0.64), whereas LSAS scores increased after PLC ($b$ = 16.20, SE = 7.40, $t$(16) = 2.19, $p$ = 0.17, $d$ = 1.72). This effect was mainly driven by the LSAS avoidance subscale (interaction of sex, treatment, and session: $F$(1,65.4) = 10.65, $p$ = 0.002) as no significant interactions of treatment with sex were observed for the anxiety subscale (all $F$ < 0.38, all $p$ > 0.54). Again, an interaction of treatment with session was significant for male participants only ($F$(1,16.2) = 9.58, $p$ = 0.007) with decreased scores after OT treatment ($b$ = -8.09, SE = 3.51, $t$(16.5) = -2.31, $p$ = 0.14, $d$ = -0.64) in contrast to increased scores after PLC ($b$ = 12.60, SE = 5.69, $t$(16.1) = 2.21, $p$ = 0.14, $d$ = 3.29).

No significant interaction of treatment with sex was evident in the female sample ($F(1,49.5) = 0.004$, $p = 0.95$). Notably, reduced social avoidance scores after OT treatment in the male sample were still evident at the follow-up measurements (T8 vs. T0: $b = -9.18$, SE = 3.21, $t(45.2) = -2.86$, $p = 0.06$, $d = -0.78$; T9 vs. T0: $b = -13.67$, SE = 3.64, $t(46.8) = -3.76$, $p = 0.005$, $d = -1.06$; all further post-hoc comparisons of the follow-up measurements: all $t < 2.77$, all $p > 0.06$), whereas still no significant interaction was found for female participants ($F(131.9) = 0.29$, $p = 0.83$).

### Supplementary tables

**Table S1**. Sociodemographic variables at study entry (T0) compared between groups

| | LL<br>(*n* = 49) | HL<br>(*n* = 78) | $\chi^2/t$ | *p* |
|---|---|---|---|---|
| Sex (female), *n* (%) | 26 (53.06) | 56 (71.79) | 3.83 | 0.05 |
| Age (years), *M* (*SD*) | 32.04 (9.62) | 34.44 (13.02) | 1.19 | 0.24 |
| Education (years), *M* (*SD*) | 18.70 (3.11) | 16.71 (3.43) | 3.38 | 0.001 |
| Monthly salary, *n* (%) | | | 13.54 | 0.04 |
| 0 to 500 € | 7 (14.29) | 9 (11.54) | | |
| 501 to 1,000 € | 11 (22.45) | 19 (24.36) | | |
| 1,001 to 1,500 € | 5 (10.20) | 7 (8.97) | | |
| 1,501 to 2,000 € | 3 (6.12) | 22 (28.21) | | |
| 2,001 to 2,500 € | 12 (24.49) | 8 (10.26) | | |
| 2,501 to 3,000 € | 5 (10.20) | 3 (3.85) | | |
| More than 3,000 € | 6 (12.24) | 10 (12.82) | | |
| BMI, *M* (*SD*) | 25.45 (3.61) | 24.90 (4.70) | 0.74 | 0.46 |
| Smoker, *n* (%) | 3 (6.12) | 11 (14.10) | 1.23 | 0.27 |
| Relationship status (single), *n* (%) | 16 (32.65) | 53 (67.95) | 13.72 | <0.001 |

*Notes.* Abbreviations: BMI, body mass index; HL, high loneliness group; LL, low loneliness group.

**Table S2**. Psychiatric symptomatology and social network description

| | **Pre-treatment (T0/T1)** | | **Post-treatment (T7)** | | | | | |
|---|---|---|---|---|---|---|---|---|
| | LL (*n* = 49) | HL (*n* = 78) | LL (*n* = 46) | HL (*n* = 70) | $t/F_{group}$ | *p* | $F_{group*session}$ | *p* |
| UCLA-L | 26.17 (3.08) | 53.70 (8.56) | 25.80 (4.22) | 51.47 (9.84) | 244.44 | <0.001 | 1.24 | 0.27 |
| WHO-5 | 17.65 (3.38) | 11.35 (4.62) | 17.48 (3.32) | 11.24 (4.90) | 52.14 | <0.001 | 0.11 | 0.74 |
| PSS-10 | 8.71 (4.75) | 19.65 (5.91) | 8.63 (4.85) | 18.61 (6.91) | 68.22 | <0.001 | 0.57 | 0.45 |
| BDI | 2.12 (3.23) | 12.83 (8.30) | 3.22 (3.48) | 13.81 (9.51) | 50.61 | <0.001 | 0.08 | 0.78 |
| LSAS total | 10.86 (11.38) | 38.69 (24.05) | 11.00 (11.45) | 40.84 (26.23) | 53.72 | <0.001 | 0.33 | 0.56 |
| LSAS anxiety | 5.94 (5.13) | 20.90 (11.24) | 6.39 (6.03) | 23.11 (13.83) | 61.43 | <0.001 | 1.58 | 0.21 |
| LSAS avoidance | 4.92 (7.91) | 17.79 (14.38) | 4.61 (6.94) | 17.73 (14.71) | 35.87 | <0.001 | 0.01 | 0.94 |
| SN social roles | 6.49 (1.77) | 4.12 (1.60) | 6.46 (2.22) | 4.66 (1.76) | 3.05 | <0.001 | 3.72 | 0.06 |
| SN people | 22.27 (8.68) | 10.63 (5.75) | 22.89 (10.02) | 12.27 (6.60) | 43.60 | <0.001 | 0.55 | 0.46 |
| SN embedded networks | 2.90 (1.37) | 1.15 (0.90) | 3.00 (1.52) | 1.39 (1.09) | 37.78 | <0.001 | 0.10 | 0.75 |
| CTQ total | 31.06 (9.51) | 45.82 (15.87) | - | - | 6.55 | <0.001 | - | - |
| CTQ emotional abuse | 6.76 (3.36) | 11.24 (5.40) | - | - | 5.77 | <0.001 | - | - |
| CTQ physical abuse | 5.76 (2.92) | 6.91 (3.30) | - | - | 2.06 | 0.04 | - | - |
| CTQ sexual abuse | 5.10 (0.47) | 6.41 (3.97) | - | - | 2.88 | 0.005 | - | - |
| CTQ emotional neglect | 7.27 (3.18) | 13.54 (5.72) | - | - | 7.93 | <0.001 | - | - |
| CTQ physical neglect | 6.18 (2.08) | 7.72 (3.11) | - | - | 3.33 | 0.001 | - | - |

*Notes:* Values are mean and standard deviation. Abbreviations: BDI, Beck Depression Inventory, Version II; CTQ, Childhood Trauma Questionnaire; HL, high loneliness group; LL, low loneliness group; LSAS, Liebowitz Social Anxiety Scale; PSS-10, Perceived Stress Scale; SN, Social Network Index; UCLA-L, UCLA Loneliness Scale; WHO-5, World Health Organization Five Well-Being Index.

**Table S3**. Sociodemographic variables at study entry (T0) compared between treatments

| | PLC<br>(*n* = 34) | OT<br>(*n* = 39) | $\chi^2/t$ | *p* |
|---|---|---|---|---|
| Sex (female), *n* (%) | 29 (85.29) | 25 (64.10) | 3.21 | 0.07 |
| Age (years), *M* (*SD*) | 36.56 (13.66) | 34.10 (12.57) | 0.80 | 0.43 |
| Education (years), *M* (*SD*) | 17.19 (2.95) | 16.41 (3.87) | 0.98 | 0.33 |
| Monthly salary, *n* (%) | | | 1.11 | 0.98 |
| 0 to 500 € | 3 (8.82) | 4 (10.26) | | |
| 501 to 1,000 € | 7 (20.59) | 9 (23.08) | | |
| 1,001 to 1,500 € | 3 (8.82) | 4 (10.26) | | |
| 1,501 to 2,000 € | 10 (29.41) | 12 (30.77) | | |
| 2,001 to 2,500 € | 4 (11.76) | 4 (10.26) | | |
| 2,501 to 3,000 € | 1 (2.94) | 2 (5.13) | | |
| More than 3,000 € | 6 (17.65) | 4 (10.26) | | |
| BMI, *M* (*SD*) | 24.71 (4.32) | 25.27 (5.11) | 0.51 | 0.61 |
| Smoker, *n* (%) | 5 (14.71) | 6 (15.38) | < 0.001 | ≈ 1 |
| Relationship status (single), *n* (%) | 23 (67.65) | 27 (69.23) | < 0.001 | ≈ 1 |

*Notes.* Abbreviations: BMI, body mass index; OT, intranasal oxytocin treatment; PLC, placebo treatment.

**Table S4**. Changes in well-being in the high loneliness group across intervention sessions

| | **Pre-treatment (T1)** | | **T2** | | **T3** | | **T4** | | **T5** | | **T6** | | **Post-treatment (T7)** | | | | | | | |
|---|---|---|---|---|---|---|---|---|---|---|---|---|---|---|---|---|---|---|---|---|
| | PLC (*n* = 30) | OT (*n* = 39) | PLC (*n* = 34) | OT (*n* = 39) | PLC (*n* = 31) | OT (*n* = 38) | PLC (*n* = 28) | OT (*n* = 34) | PLC (*n* = 22)[a] | OT (*n* = 34) | PLC (*n* = 30) | OT (*n* = 33) | PLC (*n* = 32) | OT (*n* = 37) | $F_{session}$ | *p* | $F_{treatment}$ | *p* | $F_{treatment*session}$ | *p* |
| WHO-5 | 12.6 (4.8) | 10.5 (4.4) | 11.3 (4.2) | 9.9 (3.8) | 11.6 (4.4) | 10.4 (4.5) | 12.2 (4.6) | 10.6 (4.4) | 12.7 (4.8) | 9.7 (4.7) | 11.0 (5.5) | 10.5 (4.3) | 12.7 (5.2) | 10.2 (4.2) | 0.80 | 0.57 | 2.59 | 0.14 | 0.64 | 0.70 |
| PSS-10 | 18.7 (5.7) | 20.7 (6.1) | 17.7 (5.9) | 20.8 (5.8) | 18.6 (6.2) | 20.0 (6.3) | 17.9 (6.1) | 17.8 (6.4) | 16.7 (5.8) | 19.1 (6.4) | 19.0 (6.4) | 17.9 (5.7) | 17.4 (6.9) | 19.9 (6.7) | 2.78 | 0.01 | 2.76 | 0.10 | 1.53 | 0.17 |

*Notes:* Values are mean and standard deviation. [a]For two of the PLC groups, T5 could not take place due to organizational reasons. Abbreviations: OT, intranasal oxytocin treatment; PLC, placebo treatment; PSS-10, Perceived Stress Scale; WHO-5, World Health Organization Five Well-Being Index.

**Table S5**. Changes in psychiatric outcomes and social network characteristics in the high loneliness group

| | **Pre-treatment (T0/T1)** | | **Post-treatment (T7)** | | **3-week follow-up (T8)** | | **3-month follow-up (T9)** | | | | | |
|---|---|---|---|---|---|---|---|---|---|---|---|---|
| | PLC (*n* = 34) | OT (*n* = 39) | PLC (*n* = 32) | OT (*n* = 37) | PLC (*n* = 32) | OT (*n* = 35) | PLC (*n* = 26) | OT (*n* = 24) | $F_{session}$ | *p* | $F_{treat\text{-}ment^{*} session}$ | *p* |
| UCLA-L | 51.6 (9.0) | 55.3 (8.4) | 47.8 (9.6) | 54.5 (9.2) | 50.8 (11.0) | 54.3 (10.7) | 48.8 (12.7) | 52.4 (10.4) | 4.03 | 0.008 | 1.60 | 0.19 |
| WHO-5 | 12.6 (4.8) | 10.5 (4.4) | 12.7 (5.2) | 10.2 (4.2) | 12.4 (5.5) | 10.9 (5.1) | 11.6 (6.2) | 10.7 (5.6) | 0.88 | 0.53 | 0.45 | 0.89 |
| PSS-10 | 18.7 (5.7) | 20.7 (6.1) | 17.4 (6.9) | 19.9 (6.7) | 18.0 (6.9) | 18.6 (7.0) | 19.3 (5.4) | 20.0 (7.6) | 1.95 | 0.05 | 1.08 | 0.38 |
| BDI | 12.9 (7.6) | 13.4 (9.0) | 12.4 (8.0) | 15.2 (10.6) | 13.2 (10.3) | 13.1 (10.0) | 13.2 (11.5) | 13.8 (9.6) | 0.16 | 0.92 | 0.69 | 0.56 |
| LSAS total | 38.6 (24.2) | 40.6 (24.9) | 44.6 (26.3) | 37.7 (26.4) | 34.0 (22.5) | 32.5 (24.0) | 39.1 (29.6) | 26.4 (21.8) | 4.03 | 0.008 | 1.57 | 0.20 |
| LSAS anxiety | 21.8 (11.3) | 20.7 (11.7) | 25.5 (14.3) | 21.2 (13.4) | 20.8 (11.5) | 18.9 (12.1) | 22.2 (14.3) | 15.7 (11.5) | 2.73 | 0.045 | 0.70 | 0.56 |
| LSAS avoid. | 16.8 (13.9) | 19.8 (15.1) | 19.2 (15.0) | 16.5 (14.7) | 13.2 (13.4) | 13.5 (13.2) | 16.9 (17.1) | 10.7 (12.0) | 4.84 | 0.003 | 2.14 | 0.10 |
| SN social roles | 4.2 (1.6) | 4.1 (1.6) | 5.0 (1.7) | 4.4 (1.8) | 4.4 (1.6) | 4.5 (1.9) | 4.4 (1.7) | 4.3 (1.9) | 2.79 | 0.04 | 1.35 | 0.26 |
| SN people | 10.9 (5.3) | 10.8 (6.3) | 12.8 (6.2) | 11.9 (7.1) | 10.3 (5.8) | 11.4 (6.8) | 11.0 (5.5) | 9.7 (4.7) | 2.70 | 0.047 | 1.00 | 0.39 |
| SN embed. networks | 1.1 (0.8) | 1.2 (1.0) | 1.5 (1.2) | 1.3 (1.0) | 1.2 (1.0) | 1.2 (1.0) | 1.2 (1.1) | 1.1 (0.9) | 1.58 | 0.19 | 0.54 | 0.66 |

*Notes:* Values are mean and standard deviation. Abbreviations: BDI, Beck Depression Inventory, Version II; LSAS, Liebowitz Social Anxiety Scale; LSAS anxiety, Liebowitz Social Anxiety Scale – anxiety subscale; LSAS avoid., Liebowitz Social Anxiety Scale – avoidance subscale; OT, intranasal oxytocin treatment; PLC, placebo treatment; PSS-10, Perceived Stress Scale; SN, Social Network Index; SN embed. networks, Social Network Index – embedded networks; UCLA-L, UCLA Loneliness Scale; WHO-5, World Health Organization Five Well-Being Index.

### Supplementary Figures

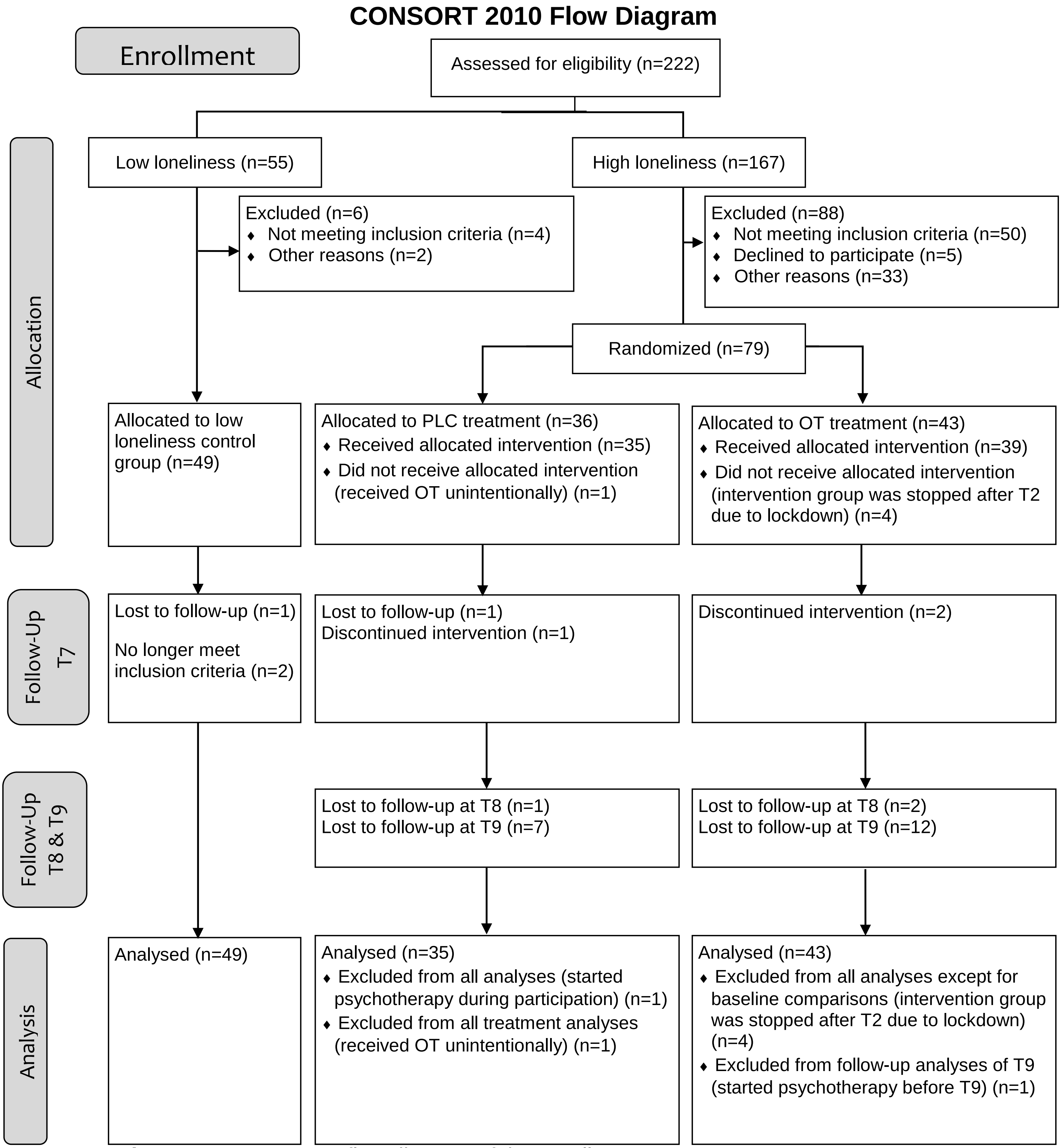


**Figure S1.** CONSORT flow diagram of the enrollment process.
